# A clustering approach to improve our understanding of the genetic and phenotypic complexity of chronic kidney disease

**DOI:** 10.1101/2023.10.12.23296926

**Authors:** A. Eoli, S. Ibing, C. Schurmann, G.N. Nadkarni, H.O. Heyne, E. Böttinger

## Abstract

Chronic kidney disease (CKD) is a complex disorder that causes a gradual loss of kidney function, affecting approximately 9.1% of the world’s population. Here, we use a soft-clustering algorithm to deconstruct its genetic heterogeneity. First, we selected 322 CKD-associated independent genetic variants from published genome-wide association studies (GWAS) and added association results for 229 traits from the GWAS catalog. We then applied nonnegative matrix factorization (NMF) to discover overlapping clusters of related traits and variants. We computed cluster-specific polygenic scores and validated each cluster with a phenome-wide association study (PheWAS) on the BioMe biobank (n=31,701). NMF identified nine clusters that reflect different aspects of CKD, with the top-weighted traits signifying areas such as kidney function, type 2 diabetes (T2D), and body weight. For most clusters, the top-weighted traits were confirmed in the PheWAS analysis. Results were found to be more significant in the cross-ancestry analysis, although significant ancestry-specific associations were also identified. While all alleles were associated with a decreased kidney function, associations with CKD-related diseases (e.g., T2D) were found only for a smaller subset of variants and differed across genetic ancestry groups. Our findings leverage genetics to gain insights into the underlying biology of CKD and investigate population-specific associations.

## Introduction

Chronic kidney disease (CKD) is a primarily asymptomatic disease characterized by a gradual loss of kidney function over a period extending from several months to years^[1]^. CKD affects approximately 9.1% of the global population, with a higher prevalence in high-income countries^[2]^. The leading risk factors for developing CKD are diabetes (40% of cases) and hypertension (29% of cases), followed by heart disease, family history of CKD, and obesity^[3]^. Other factors, such as exposure to HIV and contaminants, are additionally important in low-income countries^[4,5]^. The genetic ancestry also plays a crucial role, with increased risk rates of kidney failure in Black/African Americans and Hispanics/Latinos compared to individuals of European ancestry^[6]^. If left untreated, CKD increases the mortality risk for individuals with cardiovascular disease (CVD) and can result in the complete loss of kidney function^[7]^. Therefore, early detection is critical for improving quality of life and life expectancy. During the early stages of CKD, cost-effective treatment options are available and can be tailored to the cause of the disease^[8]^.

CKD is defined by a reduced functionality of the kidneys, which limits its filtering capability over a period of at least three months^[9]^. The main biomarkers for CKD detection include the urinary albumin/creatinine ratio (ACR) and the estimated glomerular filtration rate (eGFR) ^[10]^. While ACR facilitated diagnosing albuminuria – an indicator of kidney damage characterized by an elevated excretion of urinary albumin – the eGFR estimates the filter volume of the glomerulus per unit of time using different biomarkers such as serum creatinine^[10]^. An abnormal kidney activity is indicated by high ACR values, reduced eGFR, or both.

Over the past few decades, many large-scale genomic studies, such as genome-wide association studies (GWAS), have successfully identified more than 500 independent genetic variants associated with reduced kidney function^[11–13]^. The association between genetic variants and various phenotypes has been studied, and the results are often shared in publicly available databases, like the GWAS Catalog^[14]^. The association of one genetic variant with multiple traits can be considered to identify secondary traits associated with a phenotype. This understanding can help elucidate potentially shared disease mechanisms, assuming that genetic variants affecting a shared pathway also have a similar impact on the associated traits.

Soft-clustering methods provide a means to reduce the genetic complexity of a heterogeneous disease while also accounting for shared disease mechanisms. In contrast to hard-clustering approaches like K-means or hierarchical clustering, soft-clustering enables the factorization of high-dimensional data by identifying overlapping clusters^[15]^. Non-negative matrix factorization (NMF) is a family of algorithms within multivariate analysis that addresses the dimensionality challenge by extracting meaningful features from a given data set^[16,17]^.

In this study, we aimed to deconstruct the heterogeneity of CKD by identifying its genetic subtypes. First, we collect all published variant-trait associations for variants associated with reduced kidney function and apply soft-clustering using NMF. We used the algorithm’s weights to calculate cluster-specific polygenic scores (cPGS) within the BioMe biobank. Finally, we use a phenome-wide association study (cPGS-PheWAS) to validate and interpret the clusters. By deconstructing the complexity of CKD, this methodology contributes new insights into the disease pathways of CDK and enhances our understanding of population-specific differences for CDK.

## Results

### NMF identified nine clusters of CKD-associated variants

The most frequent CKD-associated secondary traits retrieved from the GWAS Catalog are related to kidney function (e.g., blood urea nitrogen, urea, uric acid, and cystatin C measurements), hemoglobin levels (e.g., hemoglobin measurements, hematocrit, and erythrocyte counts), T2D, body weight (e.g., body height, appendicular lean mass, BMI, BMI-adjusted waist-hip ratio), and pulse pressure (systolic and diastolic blood pressure measurements), among others (see Fig.S1). CKD-associated traits and their associated CKD variants were factorized into nine partly overlapping clusters by conducting NMF. To ensure the results were robust, we repeated the clustering with bNMF and got comparable results (Tab.S1). The top seven traits per cluster are summarised in Fig.1. The ‘Reduced lipids’ cluster is associated with decreasing blood lipid levels (triglycerides, total cholesterol, use of lipid-lowering medications) and liver enzymes. The top traits of the cluster ‘Increased body mass’ show a positive association with body weight (appendicular lean mass, body height, and body weight). The clusters ‘Increased blood volume’ and ‘Reduced blood volume’ are positively and negatively associated with volemic traits (e.g., mean corpuscular volume and mean corpuscular hemoglobin), respectively. Similarly, clusters ‘Increased/Reduced hematocrit’ show opposite associations with hemoglobin content (e.g., hematocrit, hemoglobin measurements, red blood cell density, erythrocyte count), and clusters ‘Increased/Reduced inflammation’ convey opposite associations with markers of inflammation (C-reactive protein) and blood lipids. Lastly, cluster ‘Increased urate’ is positively associated with kidney function biomarkers like urate, blood/serum urea nitrogen, blood proteins, and Cystatin C. The complete lists of the top features and variants per cluster, defined as traits and variants in the top decile of the cluster weights of the matrices H and W, are listed in Tab.S2. The matrices H and W are also available as supplementary material. Fig.S2 summarises how the variants are distributed in each cluster, showing their overlaps.

**Figure 1.**
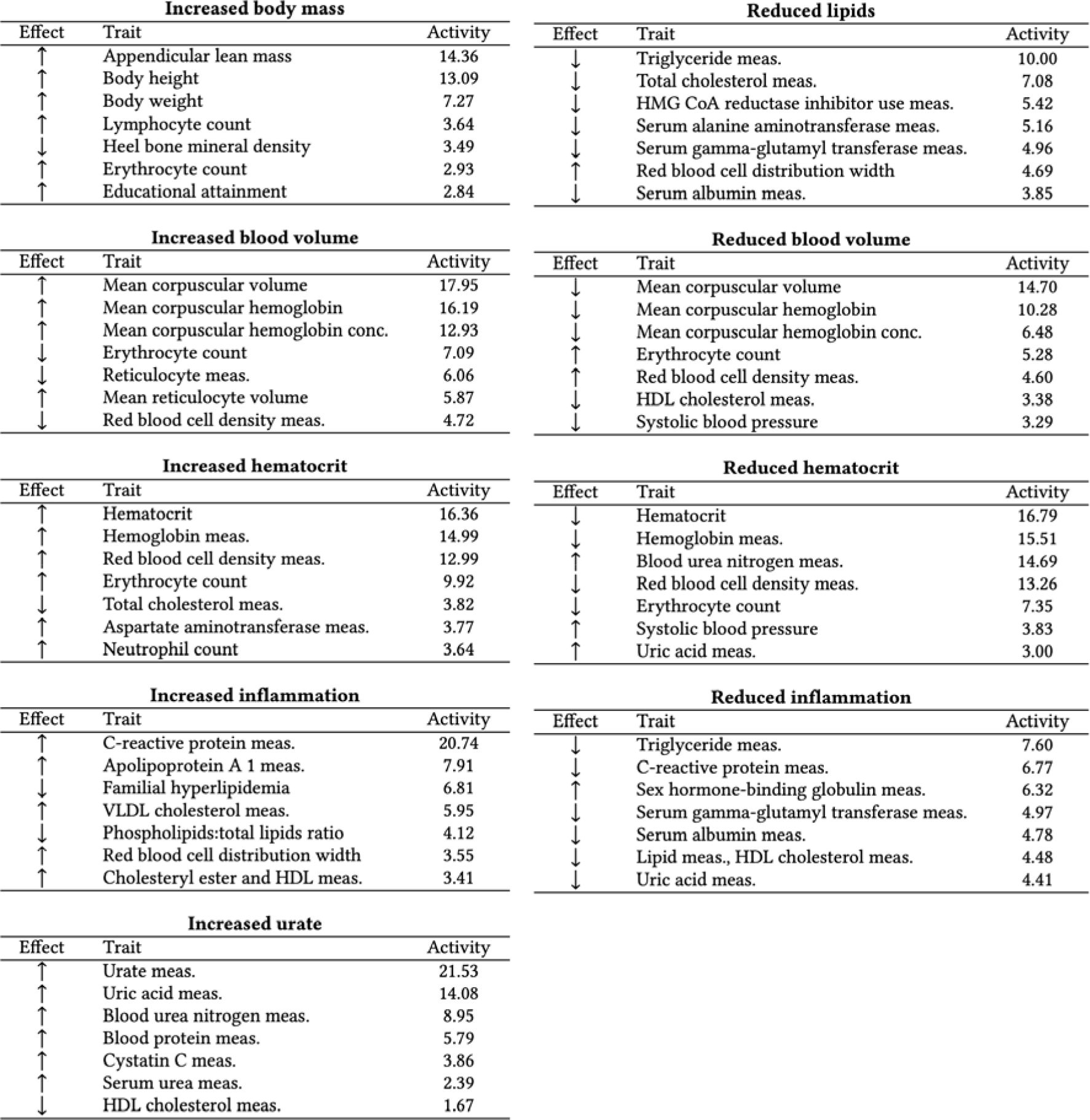
Top seven CKD-associated secondary traits per cluster. (also available as LaTeX code) The top seven secondary traits per cluster are shown with their effect direction (Effect columns) and respective cluster weights (Activity columns). ‘HDL’ is high-density lipoprotein, ‘VLDL’ is very low-density lipoprotein, ‘meas.’ is measurement, and ‘conc.’ is concentration.

### PheWAS replicated biological pathways of most clusters across ancestries

Each cluster was examined by conducting a cPGS-PheWAS with 988 quantitative traits and 832 binary traits on the four Bio*Me* cohorts (ALL, AFR, AMR, EUR). Except for the ‘Increased body mass’ and the ‘Reduced blood volume’ clusters, we replicated at least 3 of the clusters’ top 5 traits (Fig.2). The level of significance was reached more frequently across ancestries (ALL) than when replicating on the individual ones (AMR, AFR, EUR) (Tab.S3). In addition to the replicated traits, significant associations with decreasing eGFR were seen in clusters ‘Increased urate’ (β=-0.04 [-0.06 - -0.03], p-value=6.7e-09) and ‘Reduced hematocrit’ (β=-0.05 [-0.07 - -0.04], p-value=4.0e-12) (Tab.S3). ‘Reduced hematocrit’ was also nominally associated with an increased risk for chronic renal failure (OR=1.11 [1.05-1.16], p-value=1.2e-04) and with the curated phenotype ‘diabetic and hypertensive CKD’ (OR=1.27 [1.11-1.46], p-value=6.6e-04). Besides showing negative associations with disorders of lipoid metabolism, cluster ‘Increased inflammation’ shows strong negative associations with Alzheimer’s disease (OR=0.60 [0.52-0.7], p-value=1.5e-11) and dementias (OR=0.77 [0.71-0.84], p-value=1.0e-09) (Fig.S3). Regarding the individual ancestries, EUR showed the strongest associations when replicating on binary traits, with an increased risk for “visual disturbances” (OR=1.51 [1.27-1.79], p-value=2.1e-06) in the cluster ‘Reduced inflammation,’ while AFR showed the strongest associations when replicating on quantitative traits, with the strongest association being for the LDL-HDL ratio (β=-0.14 [-0.17 - -0.11], p-value=9.3e-21) in the ‘Increased inflammation’ cluster. Fig.2 summarizes which of the top traits of each cluster have been replicated, while the complete list of cPGS-PheWAS results by ancestry is stored in Tab.S3.

**Figure 2.**
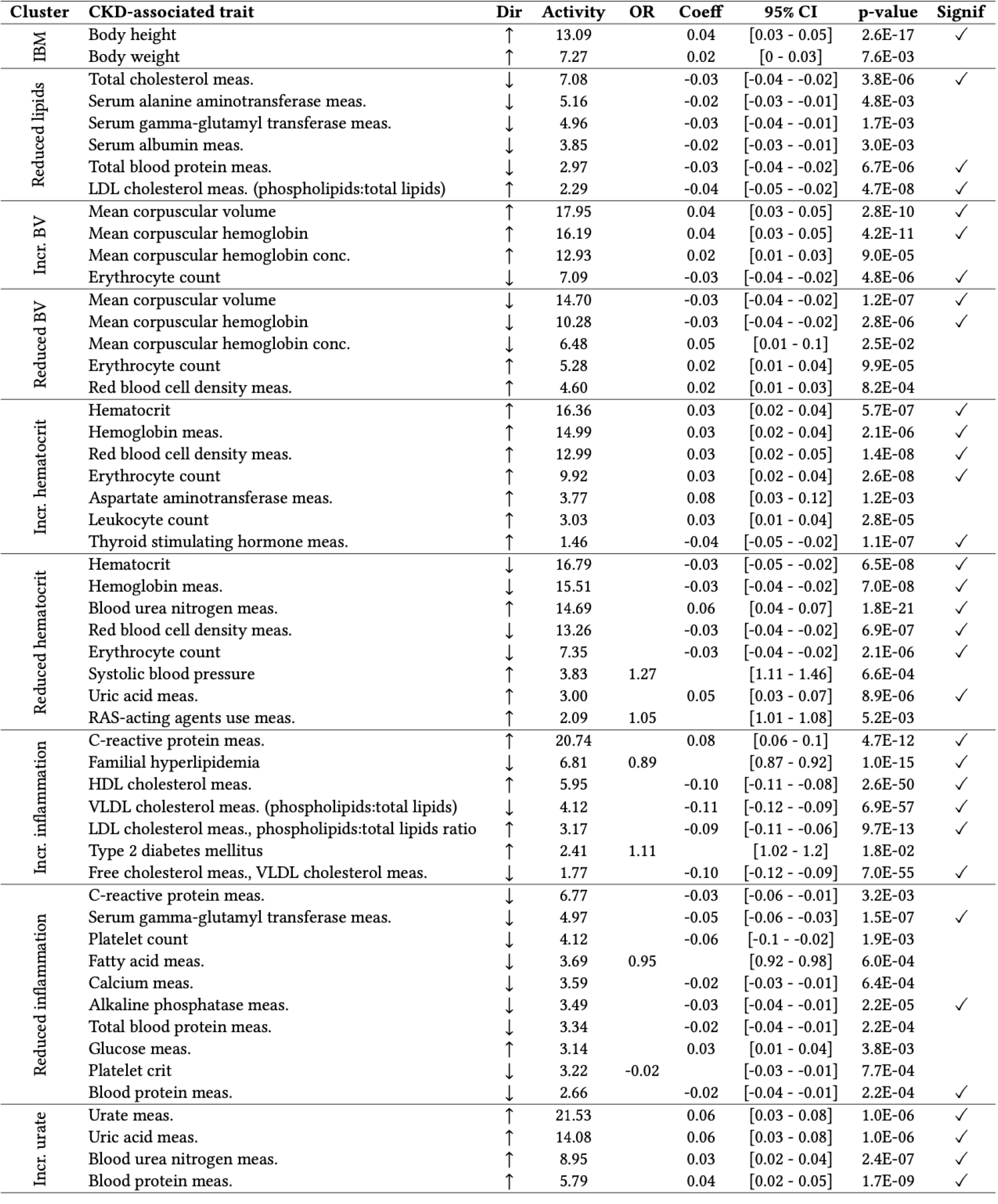
Replication of cluster traits. (also available as LaTeX code) The table lists traits replicated with the PheWAS on the ALL cohort for each cluster. ‘Dir’ is the trait effect direction, ‘activity’ is the trait cluster weight, ‘OR’ is the standardized odds ratio (binary traits cPGS-PheWAS), ‘Coeff’ is the standardized coefficient estimate (quantitative traits cPGS-PheWAS), ‘95% CI’ are the 95% confidence intervals. The last column specifies whether the p-value reaches the Bonferroni significance level. ‘HDL’ is high-density lipoprotein, ‘VLDL’ is very low-density lipoprotein, ‘RAS’ is the renin-angiotensin system, ‘meas.’ is measurement, and ‘conc.’ is concentration. Regarding the cluster names, IBM is ‘increased body mass,’ and BV is the short version for ‘blood volume.’

### cPGSs suggest clear differences between genetic ancestries

We extracted the cluster weights of the W matrix and used them to calculate cluster-specific polygenic scores (cPGS) for participants of the Bio*Me* cohort. Fig.4 shows the standardized polygenic score distributions for all NMF clusters across the Bio*Me* cohort (ALL) and the individual continental populations EUR (n=7,447), AMR (n=5,336), and AFR (n=5,660). A normal distribution was observed for the cluster ‘Increased urate’ (EUR, AMR, and AFR; Anderson-Darling test, all p-values available in Tab.S5). Although polygenic scores are expected to have a normal distribution^[18]^, the other eight clusters present either a skewed tail (e.g., ‘Increased hematocrit’) or several peaks in their cPGS distributions (e.g., ‘Reduced inflammation’). As illustrated in Fig.3, the peaks are caused by a few variants with relatively high cluster weights (the complete list of cluster weights for the top variants of each cluster is available in Tab.S2). For example, the top variant in cluster ‘Increased inflammation’ (rs429358, mapped gene: APOE) weighs 4.6, while the second one (rs17050272) weighs 0.2. In Fig.4, we can also observe how this variant is more frequent in participants of inferred EUR ancestry. Similarly, the top variant of ‘Reduced inflammation’ (rs1260326, mapped gene: GCKR) weighs 5.7 and seems to be more frequent in the AFR population, while the second one (rs4418728) weighs 0.9. This unbalance in weight creates the three peaks of the distributions: the lower peak includes the scores of individuals without the top variant (0 copies), the middle one the heterozygous (1 copy), and the higher peak includes scores of participants with two copies of the top variant. Other ancestry-specific differences are visible in the distributions of four clusters and are significant when testing with the Mann-Whitney test (all p-values available in Tab.S4). This suggests that some variants appear with different frequencies in people that do not share similar ancestry: ‘Increased inflammation’ (all combinations), ‘Reduced inflammation’ (all combinations), ‘Reduced lipids’ (EUR vs. AFR), and ‘Increased body mass’ (EUR vs AFR and AFR vs AMR).

**Figure 3.**
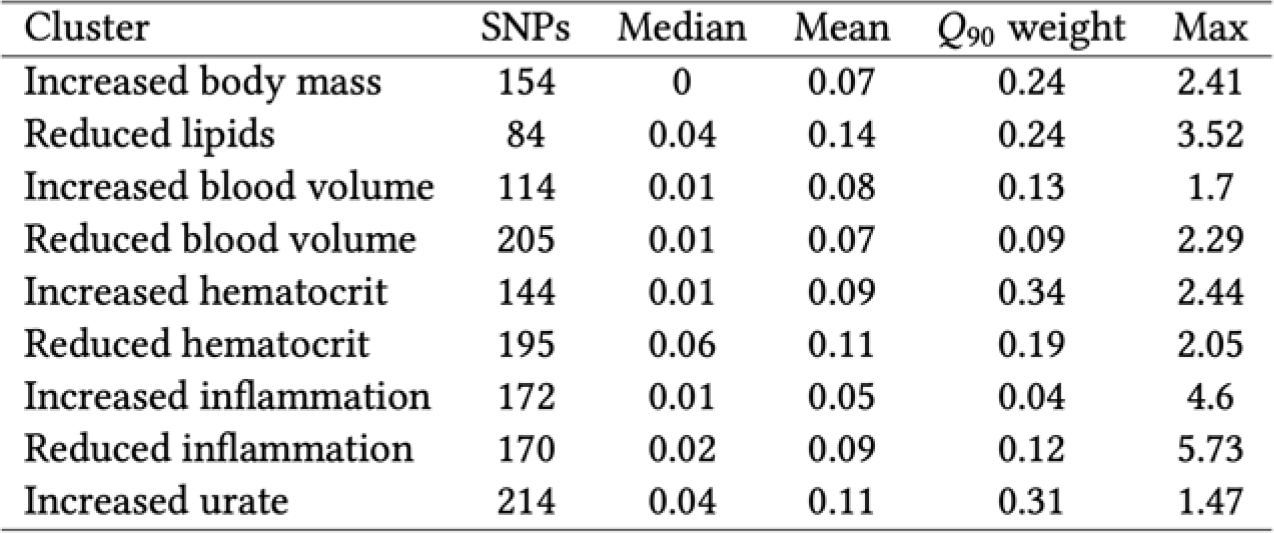
Summary statistics of the weights of each cluster. (also available as LaTeX code) ‘SNPs’ indicates the number of CKD variants with a weight > 0. The minimum weight in all clusters is 1e-45. ‘Q_90_ weight’ is the minimum weight of the SNPs in the cluster’s top decile.

**Figure 4.**
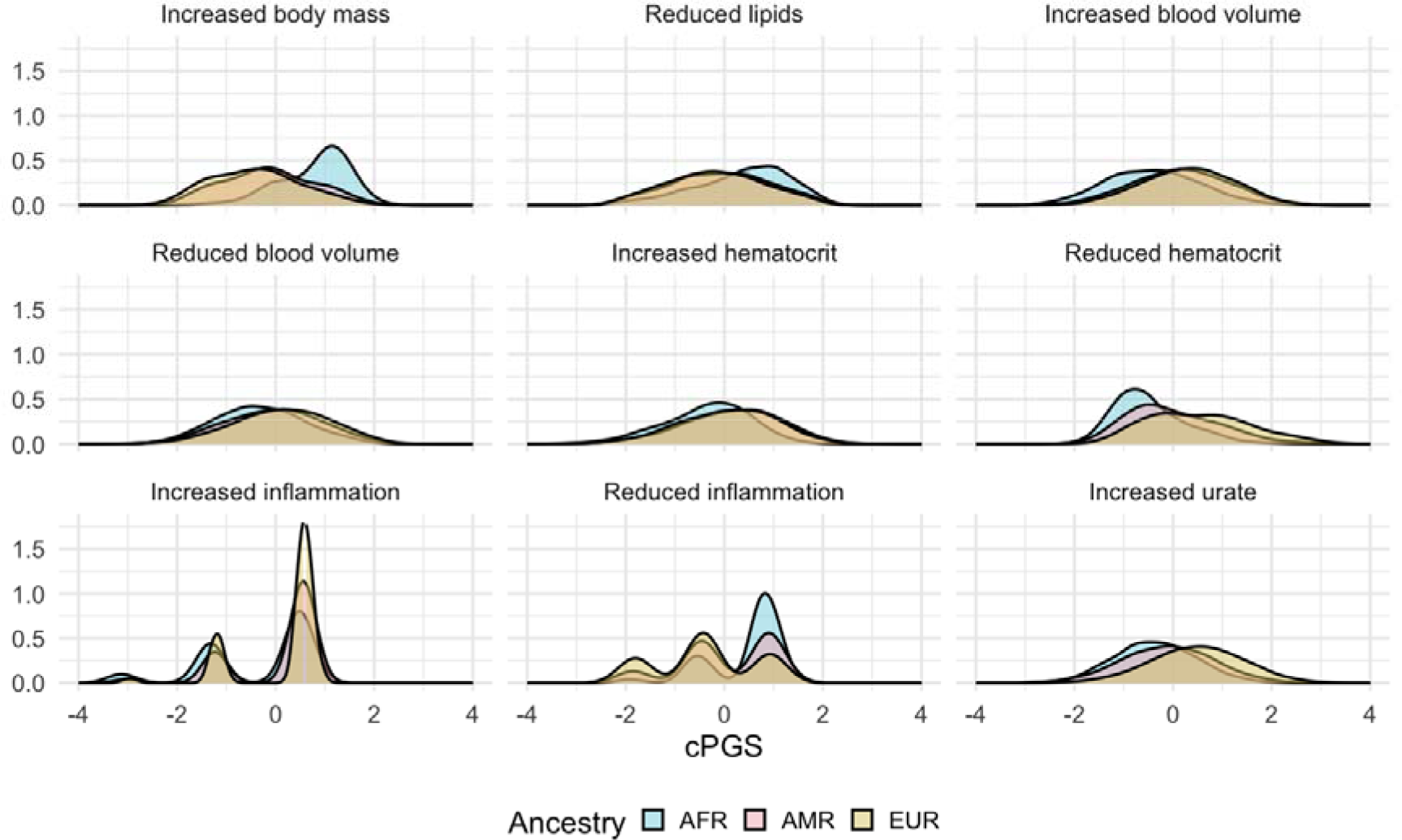
Standardized cluster-specific polygenic scores (cPGS) per genetic population. The figure compares the standardized cPGS distributions between inferred ancestries of the Bio*Me* participants. The x-axis represents the units of standard deviation (or z-scores). AFR, AMR, and EUR refer to the sub-cohorts of individuals with inferred African, Ad Mixed American, and European ancestry, respectively.

### Distribution of participants across clusters

As the cPGS are calculated separately per cluster, each Bio*Me* participant might have high polygenic scores in multiple clusters. Therefore, to understand the cluster overlap in terms of relative risk, we checked how many individuals belonged to the top decile of 1 or more clusters. 58% (18,431/31,701) of the whole Bio*Me* cohort (ALL) were at high risk in at least 1 cluster. Of these, 60.2% were in the top decile for only 1 cluster, while 37% were at risk for 2-3 clusters (Fig.S4).

## Discussion

CKD is typically defined as a progressive loss of kidney function over time. Although numerous genetic variants have been identified as associated with CKD, their relationship to disease pathways remains largely unclear. The work described here is the most comprehensive assessment of how variants associated with CKD can be grouped according to different CKD-related factors. Specifically, we included variant-trait associations of 322 CKD SNPs and 229 related metabolic traits from publicly available GWAS datasets. By analyzing these associations with NMF, a factorization approach that allows for minimal overlap between groups, we identified 9 clusters of CKD variants and associated traits.

CKD is commonly recognized as a heterogeneous condition with various underlying causes and risk factors, which are unlikely to represent a single disease process. This complexity is also reflected by the associated traits retrieved from published GWAS, which are related to kidney function, hemoglobin levels, T2D, body weight, and pulse pressure, among others. Attempting to deconvolute CKD’s genetic heterogeneity and differentially grouping these traits, the nine clusters we identified represented different aspects of CKD. For example, the ‘Increased urate’ cluster, whose clustering weights represent abnormal levels of urinary metabolites like urate, blood/serum urea nitrogen, blood proteins, and Cystatin C, is related to decreasing kidney function. In normal conditions, such blood metabolites are excreted by the kidneys, but in CKD they accumulate and exert a detrimental biological activity^[19,20]^. A second cluster, which we summarised as ‘Increased inflammation,’ was strongly clustered around rising serum C-reactive protein (CRP) concentrations. CRP is a common inflammatory biomarker in chronic diseases like CKD, diabetes, and cardiovascular diseases^[21–23]^. In line with that, patients with CKD commonly experience chronic inflammatory states^[24]^. These states tend to worsen as the disease progresses toward end-stage renal disease and are reflected, or even modulated^[25]^, by increasing CRP levels^[26–28]^.

We then studied the genotype-phenotype correlation to demonstrate the utility of the clusters. We could replicate most of the top-weighted features on quantitative traits (i.e., biomarkers), while the validation on binary traits (i.e., diagnoses) was less robust and required additional clinical interpretation. For the clusters of ‘Increased urate’ and ‘Increased inflammation,’ the top traits were confirmed by the PheWAS. CKD is also associated with dyslipidemia comprising high levels of triglycerides and LDL-cholesterol, and low levels of HDL-cholesterol and apolipoprotein A1^[29]^. We could observe similar associations in clusters ‘Increased inflammation,’ ‘Reduced lipids,’ and ‘Reduced inflammation.’ Notably, we found multiple significant associations for cluster ‘Increased inflammation’ with reduced risk of dementias. Glycerophospholipids play an essential role in neural membranes^[30,31]^, and their levels are directly correlated with serum triglycerides and inversely correlated with total cholesterol and eGFR^[32]^.

A limitation of this study is the need for more genetic diversity in the GWAS Catalog, which mainly consists of studies performed on the European population. This European bias is well described in the literature and has important implications for disease risk prediction across global populations^[33]^. Despite this lack of genetic diversity, we could still validate our results in Bio*Me*, a biobank enriched for populations with non-European ancestries. We were most powered when jointly analyzing across ancestries (ALL), while signals replicated in different ancestral groups with some group-specific differences. This result suggests that, although most CKD risk factors converge across ancestral groups, ancestry-specific studies are essential. Another two limitations are the filtering rules used to select traits and variants for the algorithm’s input matrix and the possible existence of non-additive interactions between risk factors that we did not consider in this study. Lastly, one of the input CKD studies, the PAGE study^[34]^, was also conducted using Bio*Me* data. However, this should not impact the results since we are not looking at CKD case/control scenarios but at CKD subtypes.

Understanding the biological pathways that lead to CKD is essential to improve clinical management. For example, some clusters group similar traits but with opposite effect directions (e.g., ‘Increased hematocrit’ and ‘Reduced hematocrit’), while others suggest potentially protective effects (e.g., against dyslipidemia in cluster ‘Increased inflammation’). This behavior might indicate that CKD can affect the same metabolic pathways differently, confirming the genetic complexity of the disease. Additionally, the clusters have a limited degree of overlap and, as each represents a specific set of variants, participants might be high risk (i.e., in the top decile of the polygenic score) for more than one cluster. This additive disease model, similar to the mutational signatures in cancer, suggests a possible interplay of genetic susceptibility to multiple disease-causing mechanisms^[35]^.

In summary, by clustering genetic variants associated with CKD, we identified clusters with distinct trait associations, likely representing mechanistic pathways involved in CKD. We confirmed the validity of these clusters phenotypically. Further clinical investigations could explore whether individuals with a common disrupted pathway also share similar complications, a comparable rate of disease progression, or a different treatment response. In the future, classifying patients with CKD using their genotype may improve care by offering a more personalized and genetically informed clinical plan.

## Methods

### Trait-variants selection

We identified and aligned the alleles of 508 independent genetic variants associated either with decreased kidney function (defined as low eGFR levels for at least three months) or with CKD (using ICD-9/10 codes) from the most recent GWAS and GWAS meta-analyses^[11,13,34,36,37]^ (Fig.5a). We then used the R package *LDlinkR* (R version 4.2.1) to retrieve all proxy SNPs in linkage disequilibrium (r^2^ >= 0.6) with the lead variants, across all available 1000G human populations^[38,39]^ and used the GWAS Catalog database to link the proxy SNPs to 805 associated traits (as of July 30^th^, 2022) ^[14]^. We excluded gender-specific GWAS and GWAS performed on less than 100 individuals. Additionally, as we are interested in secondary features associated with CKD, we excluded GWAS of traits directly related to eGFR or CKD (e.g., “Mild to moderate chronic kidney disease,” “Estimated Glomerular Filtration Rate”). We kept trait-variant associations with a significance threshold of less than 1×10^−6^ using a Bonferroni correction for all 2,401 associations in our data set. To reduce sparsity in the data, we excluded traits associated with less than five variants; this threshold was empirically defined by comparing the clustering results of traits associated with up to 15 CKD variants. We standardized effect sizes across all GWAS by dividing the regression coefficient beta (B) by the standard error, using the GWAS summary statistic results. Traits and variants were then arranged as a matrix with the standardized effect sizes (β) as values. Tab.S5 contains, for each input CKD variant, the list of CKD-associated secondary traits extracted from the GWAS Catalog and the corresponding exclusion criteria for those excluded during the filtering steps.

**Figure 5.**
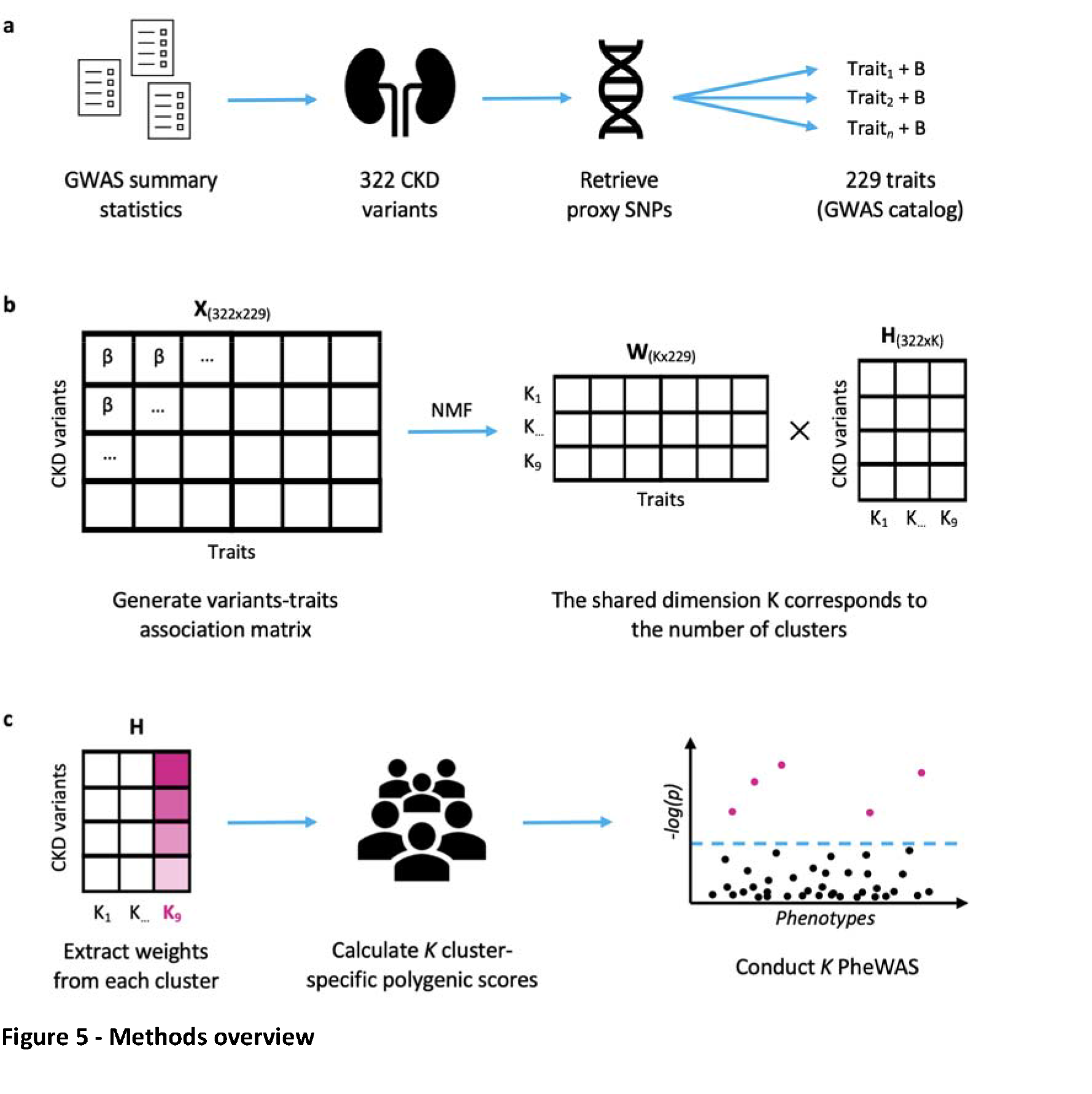
Methods overview. **a**, We selected 322 independent CKD-associated variants from the summary statistics of published GWAS. For each of them, we retrieved all independent proxy SNPs in linkage disequilibrium (r^2^ >= 0.6) and (from the GWAS Catalog) 229 proxy-associated traits with their respective effect size (B). **b**, We standardised the effect sizes across all GWAS (β) and generated an association matrix X of dimensions 229×322. NMF factorizes X into a matrix of traits (W) and one of variants (H), which share a dimension K (i.e., the number of clusters). c, We extracted the weights of each cluster from the H matrix and used them to calculate cluster-specific polygenic scores (cPGS) of 31.701 Bio*Me* participants. After standardizing the cPGS, we conducted a cPGS-PheWAS for each cluster to validate their respective top traits, which were extracted from the W matrix.

### NMF

NMF factorizes the input matrix of trait-variant associations (X, of dimensions 229×322) into a matrix of traits (H, 229xK) and one of variants (W, Kx322), so that HxW ≈ X^[16]^ (Fig.5b). The factorization rank K corresponds to the number of clusters. We implemented NMF using the R package *ButchR* with 10,000 iterations, 30 random initiations, and the convolution threshold set to 80^[40]^. The number of expected clusters was set between 2 and 20. ButchR suggests the optimal K based on six cluster evaluation metrics, like the mean silhouette width and the Frobenius error. If two or more K were presented, we considered results with the highest mean silhouette width and the lowest Frobenius error, as suggested by Alexandrov et al^[35]^. As additional validation, we also performed a Bayesian version of NMF^[17]^, using the code provided by Udler et al. bNMF was run 1,000 times with up to 200,000 iterations in each run.

### Cluster-specific polygenic scores

The results of clustering provide cluster-specific weights for each variant and trait. We used *PLINK* and the variant cluster weights to calculate cluster-specific polygenic scores (cPGS) of the Bio*Me* biobank participants^[42]^. cPGS were standardized within each cluster. The normality of each cPGS distribution was tested with the Anderson-Darling method. Differences between ancestry-specific distributions were tested with the Mann-Whitney test.

### Validation cohort (Bio*Me*)

We validated our results using the genetic and linked electronic health records (EHR) data of 31,701 Bio*Me* biobank participants^[43]^ (Fig.5c). As a fine-scale population structure can improve the risk prediction of complex diseases within genetic groups^[44]^, we inferred the genetic ancestry of the Bio*Me* participants. We then performed a Principal Component Analysis (PCA) using *PLINK*, excluding relatives above 2nd-degree (kinship method, estimated using *KING*^[45]^) and variants with minor allele frequency below 0.05^[42,46]^. We trained a random forest classifier to infer the genetic ancestry of Bio*Me* participants using the 1000 Genomes labels as reference^[47]^. The labeled ancestries are Admixed American (AMR, n=5,336), African (AFR, n=5,660), European (EUR, n=7,447), South Asian (SAS, n=613), and East Asian (EAS, n=728). For sub-population-specific analyses, we removed participants with mixed ancestry (defined as having a random forest probability ≤ 0.5) and outliers by only including the quantiles 0.25-0.90^[48]^ (n=11,404).

### Modeling disease outcomes as a function of cluster-specific polygenic scores

For each cluster, the cPGS were associated with the phenotypes available in the Bio*Me* data set by performing a phenome-wide association study (cPGS-PheWAS). We fitted linear regression models to analyze 988 quantitative traits (e.g., laboratory results) and logistic regression models for 832 binary traits with cPGS as independent variables, adjusting for sex, age, and the first ten genetic principal components (*stats* R package^[49]^). Binary traits included Phecodes mapped to ICD-9 and ICD-10 codes (a Phecode is considered if at least two relevant diagnostic codes were present in a patient’s EHR)^[50]^ and curated phenotypes^[51]^. Controls were identified as the reference category. Traits were only considered if present or measured in at least 100 biobank participants. The model parameters were standardized using the *effectsize* R package (refit method) ^[52]^. Standardized coefficient estimates (linear regression) and odd ratios (logistic regression) were reported with the corresponding 95% confidence intervals. The Bonferroni method was used to adjust for multiple testing, and the alpha threshold was defined as 2.7e-05 (0.05/(988+832)). We then compared the PheWAS results with the traits in the top decile of NMF’s trait weights.

## Acknowledgment

We would like to thank Miriam Udler, who kindly provided us with the script of the bNMF algorithm, which was used in this study to confirm the clusters identified by NMF. This work was supported in part through the computational and data resources and staff expertise provided by Scientific Computing and Data at the Icahn School of Medicine at Mount Sinai and supported by the Clinical and Translational Science Awards (CTSA) grant UL1TR004419 from the National Center for Advancing Translational Sciences. Additionally, this work was supported by the Office of Research Infrastructure of the National Institutes of Health under award number S10OD026880, which allowed us to use Mount Sinai Data Warehouse (MSDW) data. Regarding HPI.MS resources, funding was provided by the Hasso Plattner Foundation (HPF). Additionally, the research leading to these results has received funding from the Horizon 2020 Programme of the European Commission under Grant Agreement No. 826117 (Smart4Health). The Mount Sinai BioMe Biobank has been supported by The Andrea and Charles Bronfman Philanthropies and in part by Federal funds from the NHLBI and NHGRI (U01HG00638001; U01HG007417; X01HL134588). We thank all participants in the Mount Sinai BioMe Biobank. We also thank all of our recruiters who have assisted in data collection and management, and we are grateful for the computational resources and staff expertise provided by Scientific Computing at the Icahn School of Medicine at Mount Sinai.

## Contributions

Conceptualization, C.S., E.B.; Methodology, C.S., S.I.; Data analysis, A.E., S.I.; Writing of the original draft, A.E.; Tables and Figures, A.E.; Supervision, S.I., H.O.H., E.B.; Project administration, E.B.; Funding acquisition, E.B., H.O.H. All authors reviewed and edited the manuscript. All authors have read, discussed, and approved the manuscript, its analyses, and interpretations.

## Data availability statement

All publicly available data (input variants, trait-variant associations) used to support the findings of this study are included in this published article (and its Supplementary Information files) and are also available from the cited publications and GWAS Catalog. Additional data generated for the analysis steps, including source code and intermediate results, are available from the corresponding author upon reasonable request. The data used to validate the findings of this study are available from BioMe biobank (https://icahn.mssm.edu/research/ipm/programs/biome-biobank), but restrictions apply to their availability. To access the data, please reach out to.

