## Supplementary Material for "A clustering approach to improve our understanding of the genetic and phenotypic complexity of chronic kidney disease"

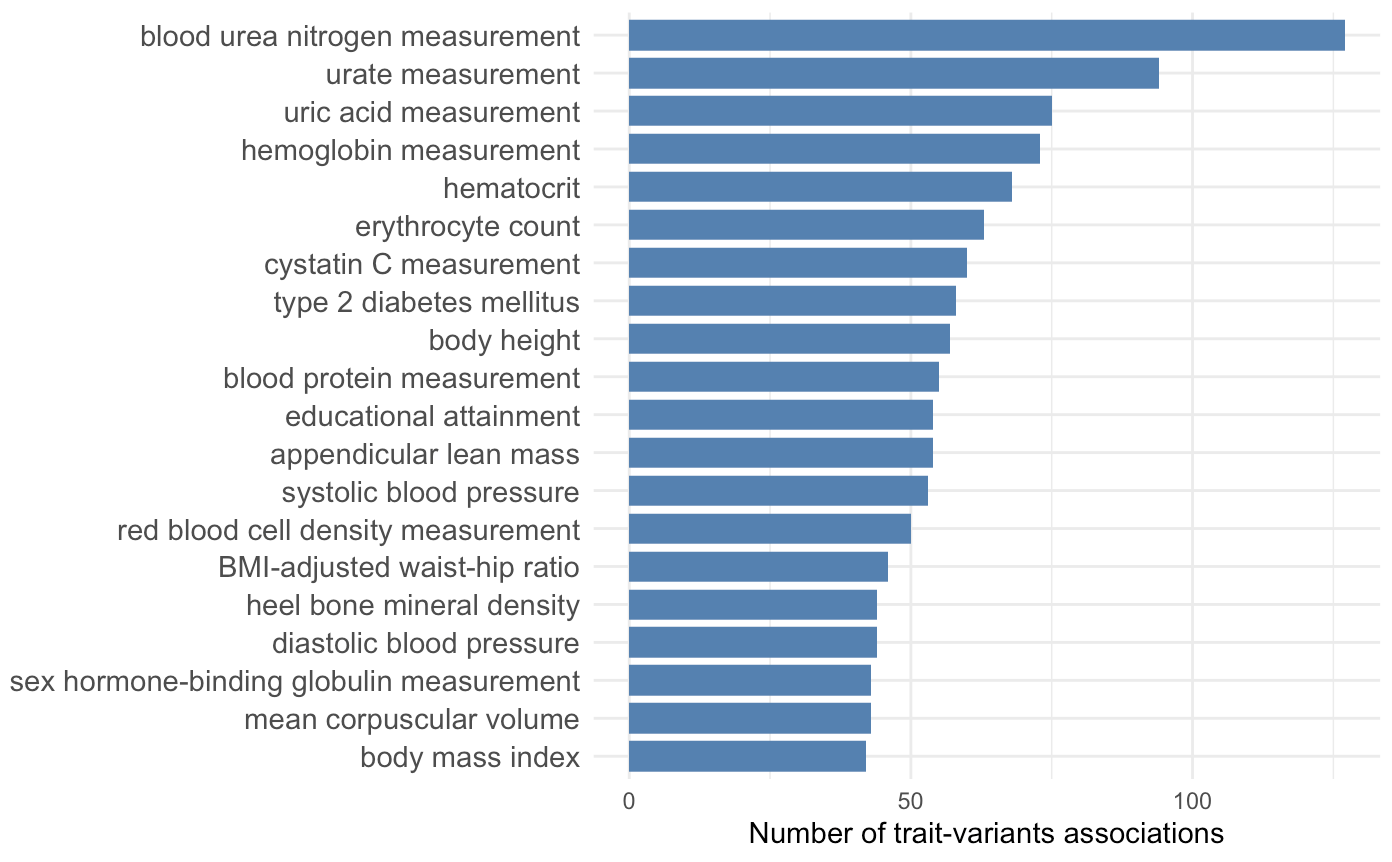

**Figure S1 - Top 20 most frequent CKD-associated traits**

Number of the most frequent traits associated with the lead CKD variants.

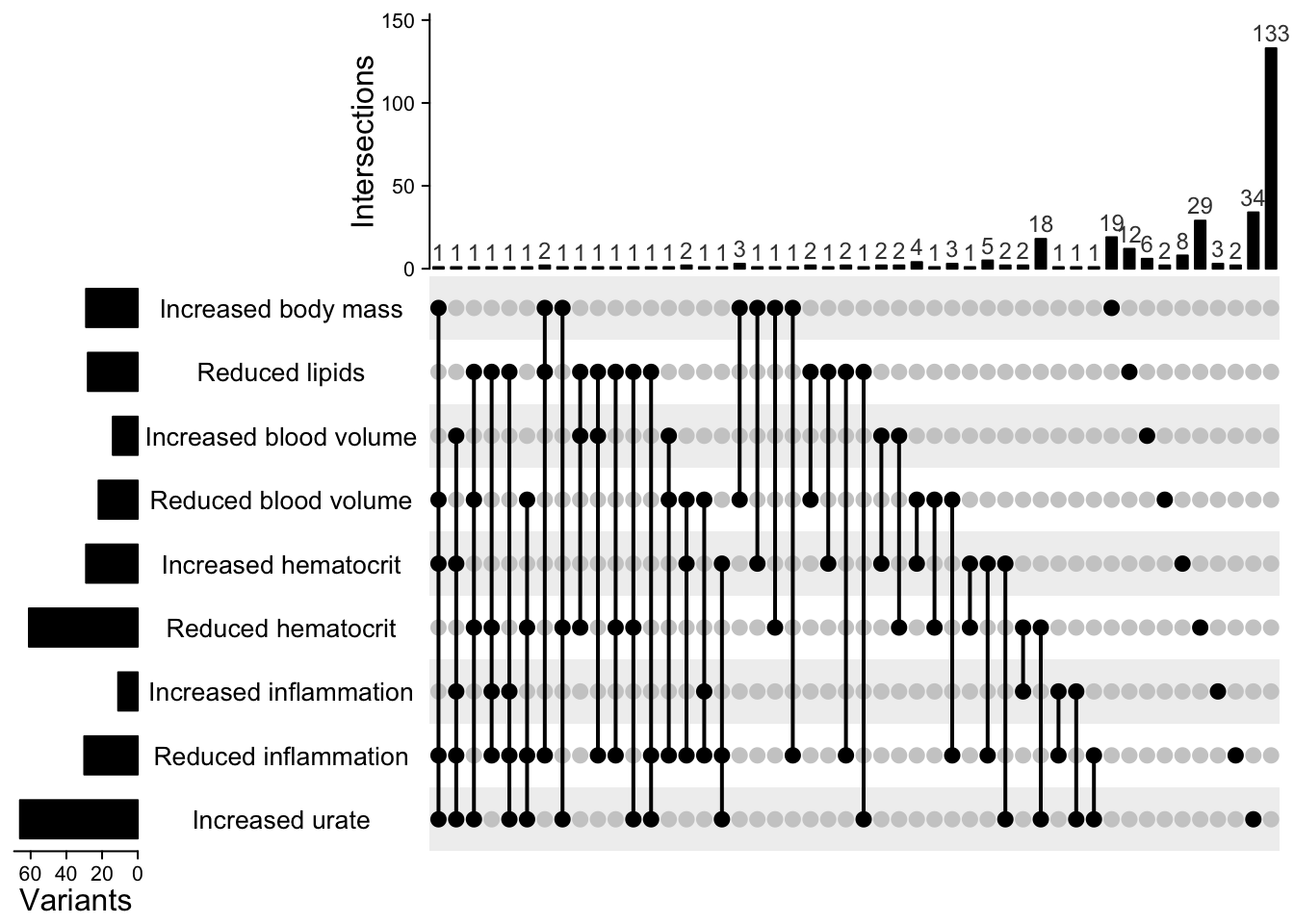

**Figure S2 - Variants overlap and distribution per cluster**

On the left, the figure shows the number of variants in each cluster. On the top, the figure shows one column per each type of cluster intersection for variants in the top decile of the overall weight distribution. The dotted lines indicate which clusters share the number of variants specified on top of the column, while single dots indicate that those variants are unique for the corresponding cluster. The last column on the right represents the number of variants not in the distribution's top decile.

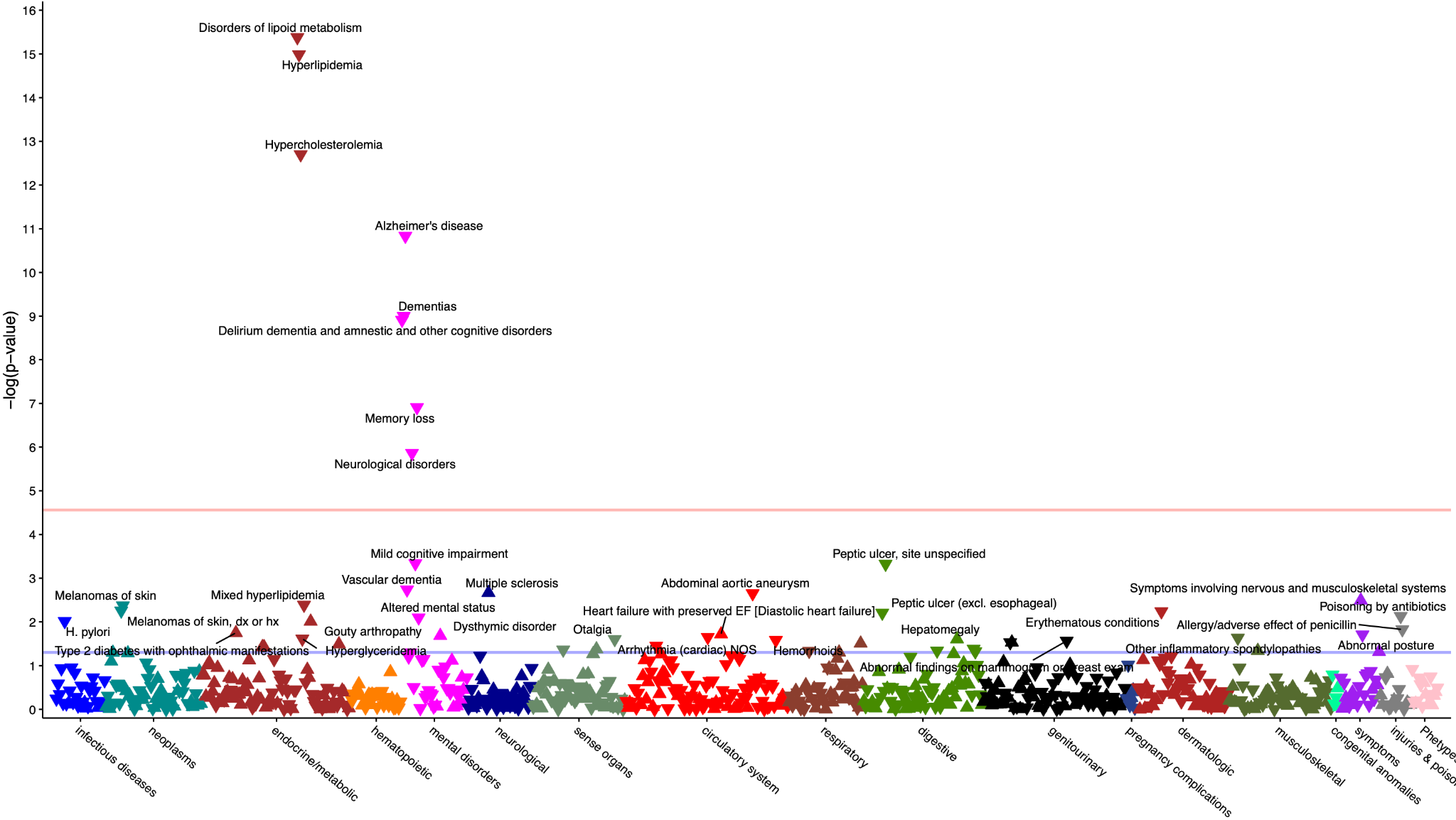

**Figure S3 - Validation of cluster ‘Increased inflammation’**

PheWAS results of cluster ‘Increased inflammation’ on binary traits. The blue line represents the nominal significance level (0.05), and the red line represents the significance level adjusted for multiple testing (2.7e-05). The labeled traits have a p-value smaller than 0.05. An upward-pointing triangle represents a positive association, while a downward-pointing triangle represents a negative one. Each color represents a group of phenotypes.

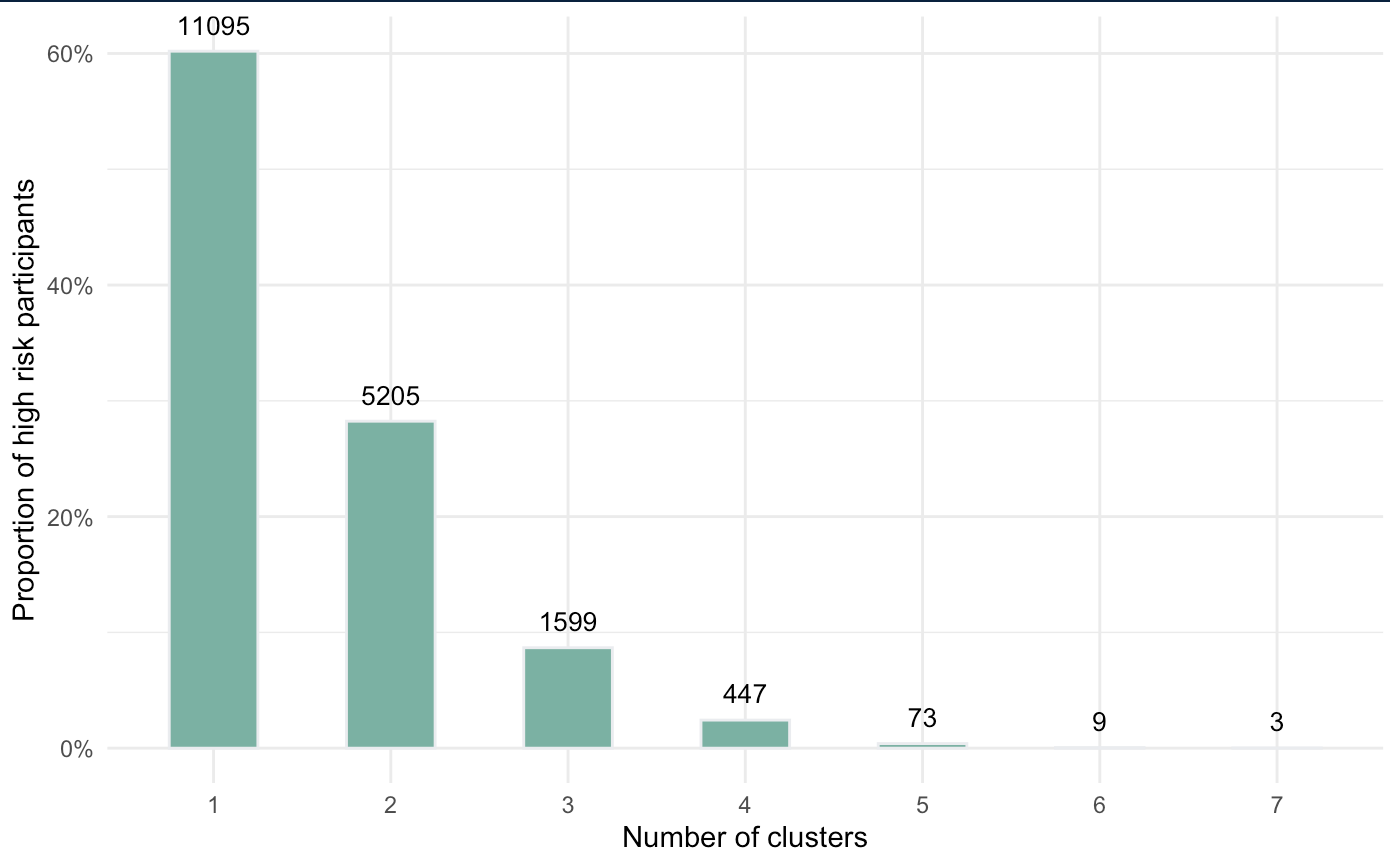

**Figure S4 - Proportion of individuals at high risk in at least one cluster**

The number on top of each bar represents the count of Bio*Me* participants in the top decile of cPGS for at least one cluster.

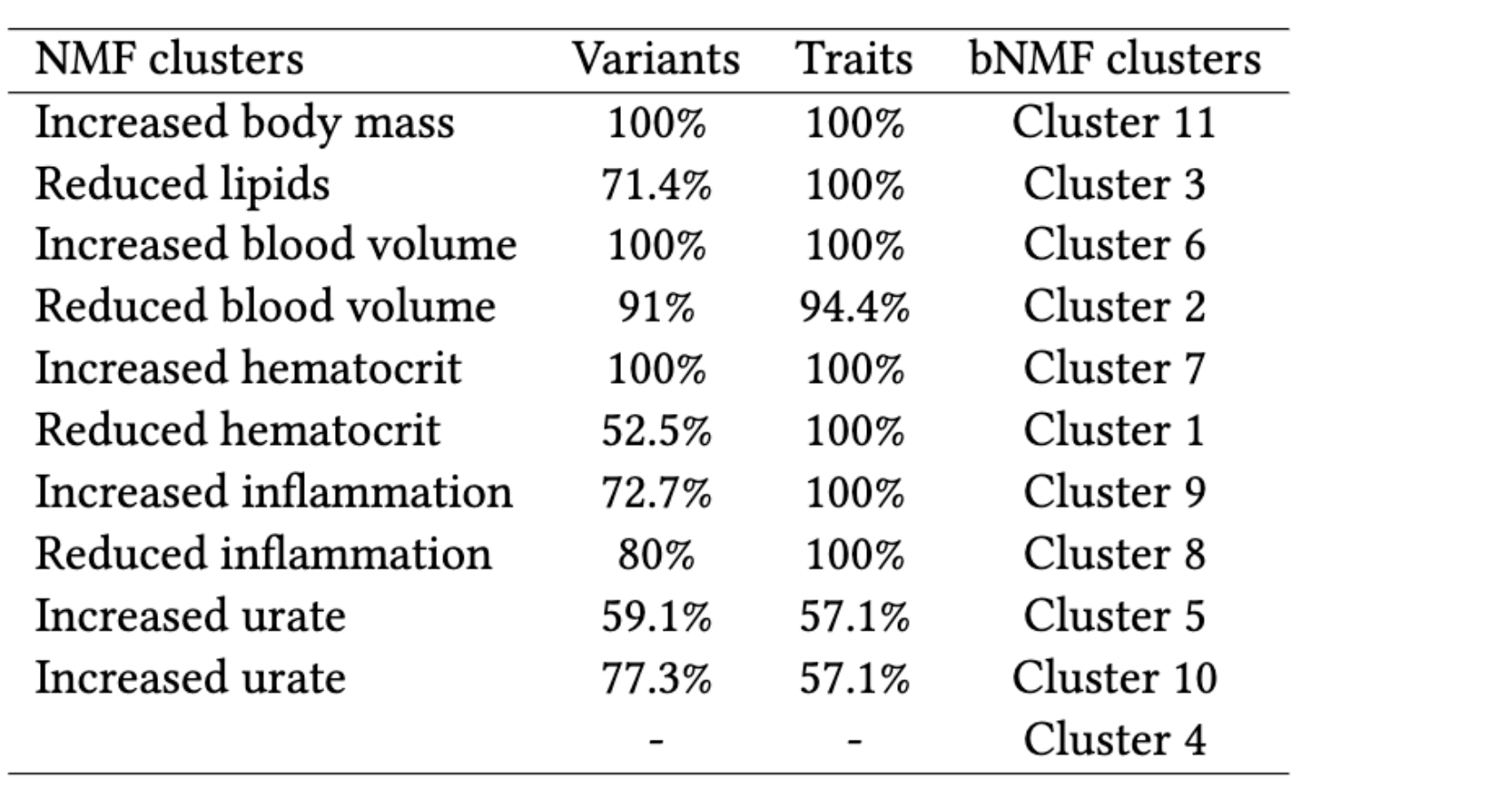

**Table S1 - Results comparison between NMF and bNMF** (also available as LaTeX code)

bNMF resulted in 11 clusters. For each NMF-bNMF cluster combination, we compared the proportion of identical variants and traits in the top decile of the cluster weights. We considered relevant only results with an overlap of variants and traits higher than 50%.

Supplementary Table S2 - Top variants and traits per cluster.xlsx

**Table S2 - Top variants and traits per cluster**

List of the top variants and traits for each cluster with the respective weights extracted from the H and W matrices of the NMF results.

Supplementary Table S3 - PheWAS replication check.xlsx

**Table S3 - Validation of clusters’ traits and cPGS distribution tests**

Sheet 1 (‘Feature validation (ALL)’) compares the top clusters’ features with the PheWAS results of binary and quantitative traits for the cohort ‘ALL.’ Sheet 2 (‘PheWAS - Quant. traits’) and sheet 3 (‘PheWAS - Binary traits’) contain the cPGS-PheWAS results of all cohorts (ALL, EUR, AFR, AMR) for quantitative and binary traits, respectively.

Supplementary Table S4 - cPGS distribution testing.xlsx

**Table S4 - cPGS distribution testing**

Sheet 1 (‘cPGS ancestries distr test’) contains the p-values of the Mann-Whitney test, which was used to test the cPGS distribution of each pair of ancestries. Sheet 2 (‘cPGS normality distr test’) contains the p-values of the Anderson-Darling test, which was used to test the normality of each cPGS distribution (by ancestry and cluster).

Supplementary Table S5 - CKD secondary traits.xlsx

**Table S5 - Full list of CKD-associated phenotypes from the GWAS Catalog**

A list of proxy SNPs is shown for each independent CKD-associated SNP. Each proxy SNP is then associated with one or more phenotypes, which were extracted from the GWAS Catalog. For each of these associations we provide: CKD SNP (rsID), Proxy SNP (rsID), Mapped trait (proxy SNP's associated trait), Date added to GWAS Catalog, PUBMED ID, Initial sample size, Replication sample size, Region, Chr, Position (hg19), Reported gene(s), Mapped gene(s), Upstream gene ID, Downstream gene ID, SNP gene IDs, Strongest SNP-Risk allele, Context, Effect, Risk Allele frequency, p-value, -log10(p-value), Effect size (OR or Beta), 95% CI (text), Genotyping technology, Alleles, Effect allele, MAF, Distance, Dprime, R2 (LD), Position (hg38), Standardised OR/Beta, CI (lower), CI (upper), SE, Exclude, Exclusion criteria.
